# Phylodynamics of SARS-CoV-2 transmissions in France, Europe and the world during 2020

**DOI:** 10.1101/2022.08.10.22278636

**Authors:** Romain Coppée, François Blanquart, Aude Jary, Valentin Leducq, Valentine Marie Ferré, Anna Maria Franco Yusti, Lena Daniel, Charlotte Charpentier, Samuel Lebourgeois, Karen Zafilaza, Vincent Calvez, Diane Descamps, Anne-Geneviève Marcelin, Benoit Visseaux, Antoine Bridier-Nahmias

## Abstract

**Background:** Although France was one of the most affected European countries by the COVID-19 pandemic in 2020, the dynamics of SARS-CoV-2 transmissions within France, Europe and worldwide remain only partially characterized during the first year of the pandemic.

**Methods:** Here, we analyzed GISAID deposited sequences from January to December 2020 (n = 638,706 sequences). To tackle the huge number of sequences without the bias of analyzing a single sequence subset, we produced 100 independent and randomly selected sequence datasets and related phylogenetic trees for different geographic scales (worldwide, European countries and French administrative regions) and time periods (first and second half of 2020). We applied a maximum likelihood discrete trait phylogeographic method to date transmission events and to estimate the geographic spread of SARS-CoV-2 to, from and within France, Europe and worldwide.

**Results:** The results unraveled two different patterns of inter- and intra-territory transmission events between the first and second half of 2020. Throughout the year, Europe was systematically associated with most of the intercontinental transmissions, for which France has played a pivotal role. SARS-CoV-2 transmissions with France were concentrated with North America and Europe (mainly Italy, Spain, United Kingdom, Belgium and Germany) during the first wave, and were limited to neighboring countries without strong intercontinental transmission during the second one. Regarding French administrative regions, the Paris area was the main source of transmissions during the first wave. But, for the second epidemic wave, it equally contributed to virus spread with Lyon and Marseille area, the two other most densely populated cities in France.

**Conclusion:** By enabling the inclusion of tens of thousands of viral sequences, this original phylogenetic strategy enabled us to robustly depict SARS-CoV-2 transmissions through France, Europe and worldwide in 2020.

## Background

On 1st December 2019, an outbreak of severe respiratory disease was identified in the city of Wuhan, China (Huang et al., 2020). Rapidly, the agent of the disease was identified as the severe acute respiratory syndrome coronavirus 2 (SARS-CoV-2) (Zhu et al., 2020), responsible for the ongoing global pandemic of coronavirus disease 2019 (COVID-19). By the end of 2020, the virus had caused over 1.8 million deaths worldwide including ∼65k deaths in France, concomitantly with social and economic devastations in many regions of the world (Mofijur et al., 2021; Santomauro et al., 2021). Since the beginning of COVID-19 pandemic, the scientific community has stepped up to thoroughly characterize the virus, including its pathogenesis, the monitoring of its circulation in human populations, and the development of several treatments or vaccines (Cevik et al., 2020; Krammer, 2020). The development of epidemiological models have been particularly helpful to evaluate the viral spread both in short- and long-terms and inform the public health decisions (Hoertel et al., 2020; Kissler et al., 2020).

In addition to clinical and epidemiological insights, the viral whole-genome sequencing has become a powerful and invaluable tool to better understand the infection dynamics (Volz et al., 2013). The number of SARS-CoV-2 whole-genome sequences available have rapidly grown thanks to an altruistic and international effort of scientist departments gathered via the Global Initiative on Sharing All Influenza Data, GISAID (https://www.gisaid.org/) (Khare et al., 2021). These genomic sequences are essential to effectively reconstruct the global viral spread and the origins of variants. Today, genomic data have become a strong asset in addition to epidemiological data to inform governments, helping to define and assess the most appropriate public health measures (Attwood et al., 2022; Rife et al., 2017). Until now, the existing phylogenetic tools cannot include large number of data such as those generated by widespread viral sequencing and the development of specific frameworks is necessary.

In France, the first COVID-19 suspected case has been identified in late December 2019 (Deslandes et al., 2020), and the first confirmed cases of SARS-CoV-2 infection were detected on the 24th January 2020, in individuals who were recently arrived from China (Bernard Stoecklin et al., 2020). COVID-19 cases remained scarce until the end of February, when the national incidence curve of new SARS-CoV-2 infections started to rise (**Figure 1**). By the end of February, reinforced measures were announced, including social distancing, cessation of passenger flights to France, school closure, and finally, a complete lockdown across the entire country from 17th March to 10th May 2020. The reported daily incidence and numbers of severe cases peaked at the beginning of April 2020 before decreasing steadily until August 2020. However, after relaxation of social distancing in June, a second wave of infections occurred in early September peaking at more than 100k positive cases in a single day on the 2nd November 2020 (**Figure 1**). As for the first wave, daily incidence and severe COVID-19 cases gradually diminished down to a number of positive daily cases varying between 2k and 25k at the end of the year after a second national lockdown between the 29th October and 15th December 2020. These epidemiological tendencies were somehow similar in most European countries except for Russia or Romania, where high rates of SARS-CoV-2-related deaths were reported even during the summer of 2020. Of note, the other continents showed different patterns of virus circulation: the number of deaths increased about two weeks later in North America; and from early May, Asia, North America and South America were highly impacted by the pandemic (**Figure 1—figure supplement 1**).

**Figure 1.**
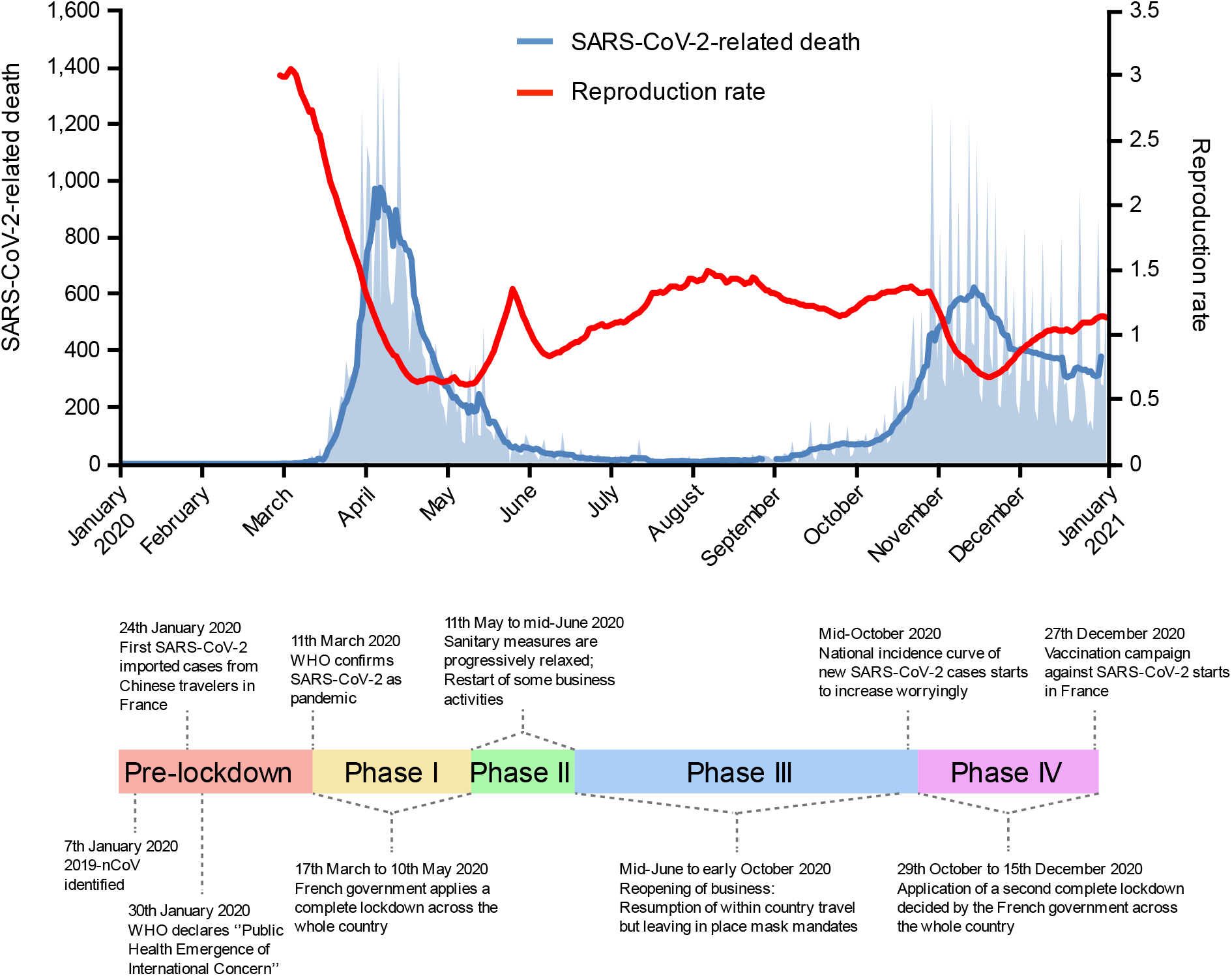
Timeline of SARS-CoV-2-related deaths and reproduction rate in France, 2020. Key events are indicated on the timeline. Official lockdowns included stay home orders and closure of schools and daycares. The two first French epidemic waves are respectively dated of March to end-June 2020, and September to December 2020. SARS-CoV-2-related deaths are displayed as the daily number of deaths (light blue area) and as the mean of deaths in a week (curve).

Elucidating the SARS-CoV-2 dynamic throughout the various phases of the pandemic is paramount to get lessons for future viral epidemics (Rife et al., 2017). Here, we analyzed GISAID deposited sequences to elucidate the origins and spread of the virus in France, Europe and worldwide from January to December 2020. Through a maximum likelihood discrete trait phylogeographic method, we estimated the main geographical areas that contributed to viral introductions in France and Europe, the countries/continents to which France transmitted SARS-CoV-2 the most and the contribution of the different French regions to the national circulation of the virus. We explored the differences in viral circulation during each of the two European epidemic waves of 2020 independently.

## Results

### Description of the datasets and global diversity of SARS-CoV-2 sequences

Prior to any phylodynamics investigation, we checked whether the sets of sequences produced for each geographic scale (i.e., worldwide, Europe and French regions) and time period (i.e., the first and second European epidemic waves) matched with the number of SARS-CoV-2 infections in 2020. This step was necessary to ensure that our sampling method did not introduce too strong biases from the start. For each geographic scale and time period, we noticed a positive correlation with the estimated SARS-CoV-2-related deaths (Spearman’s rank correlation, *p* < 0.001; r = 0.94 for the lowest correlation). We also confirmed that the number of sequences per territory was, on average, properly temporally distributed within each time period (**Figure 2**).

**Figure 2.**
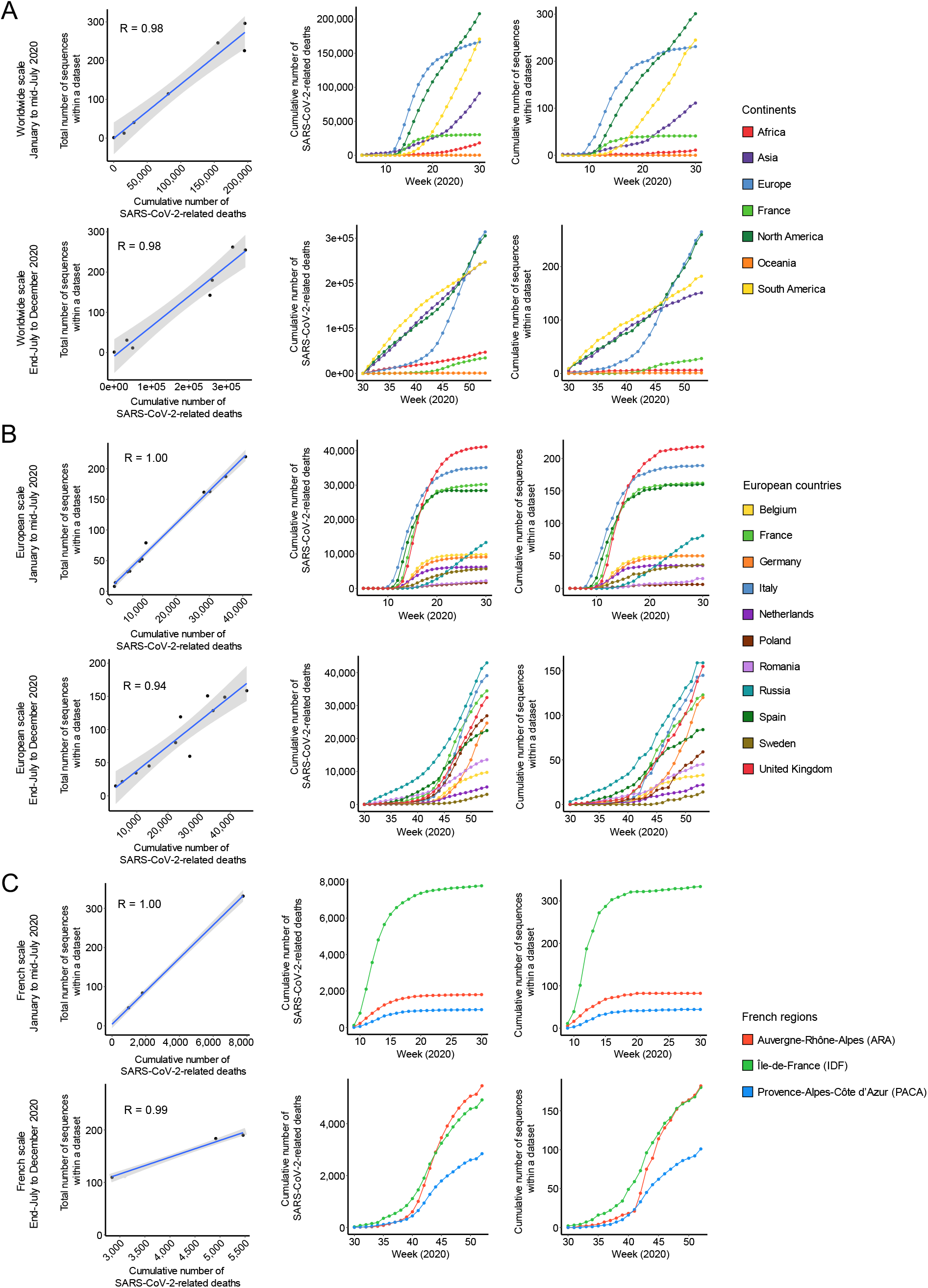
Positive correlation between the number of SARS-CoV-2-related deaths and the number of sequences included within a dataset at (A) worldwide, (B) European and (C) French scales. For each item, the first row of plots corresponds to the data from January to mid-July 2020, and the second one covers the period of end-July to December 2020. The *left* plots show the positive correlation between the total number of sequences per territory and the number of SARS-CoV-2-related deaths during the period investigated. The coefficient of correlation R is indicated inside the plots. The *central* and *right* plots show the cumulative number of SARS-CoV-2-related deaths and sequences within a dataset over time. Each color corresponds to a territory.

Some countries and French administrative regions were however discarded in the analyses because they were not sufficiently represented in the GISAID database. Overall, a total of 41,566 and 43,368 distinct SARS-CoV-2 sequences were included across the 100 phylogenies for the worldwide dataset, respectively for the first and the second time periods investigated (**Table 1**). At the European scale, 28,235 and 29,369 different SARS-CoV-2 sequences covering eleven countries were analyzed across the 100 replicates (**Table 1**). Focusing on French administrative regions, sequences available on the GISAID database were very sparse. The Provence-Alpes-Côte d’zur (PACA, Southeast of France, Marseille area) was the only region that highly sequenced SARS-CoV-2 in 2020. Île-de-France (center of France, Paris area) and Auvergne-Rhône-Alpes (ARA, East of France, Lyon area) have sequenced much less than PACA, but provided sufficient data to investigate SARS-CoV-2 transmission events within France. The remaining French administrative regions were discarded since no sufficient sequences were available to properly match the number of weekly deaths (**Figure 2—figure supplement 1**). We thus considered 2,810 unique sequences across the 100 replicates for the first half of 2020, and 3,639 unique sequences for the second time period studied (**Table 1**).

**Table 1.** Number of SARS-CoV-2 sequences investigated within each dataset.

| Dataset | Geographical categories | Period investigated | Average number of sequences within a set | Number of sequences investigated |
| --- | --- | --- | --- | --- |
| World | Africa, Asia, Europe, France, North America, Oceania, South America | January to mid-July 2020 | 935 | 41 566 |
|  |  | End-July to December 2020 | 882 | 43 368 |
| Europe | Belgium, France, Germany, Italy, The Netherlands, Poland, Romania, Russia, Spain, Sweden, United Kingdom | January to mid-July 2020 | 999 | 28 235 |
|  |  | End-July to December 2020 | 961 | 29 369 |
| France | Auvergne-Rhône-Alpes (ARA), Île-de-France (IDF), Provence-Alpes-Côte d'Azur (PACA) | January to mid-July 2020 | 462 | 2 810 |
|  |  | End-July to December 2020 | 483 | 3 639 |

The genomic diversity of circulating SARS-CoV-2 in the different continents, countries and French regions was found to be globally uniform (**Figure 2—figure supplement 2**). Overall, genomes showed high sequence conservation compared to the Wuhan-Hu-1 reference in 2020 (mean and median of ∼13 SNPs with 95% of the distribution comprised between 4 and 25 SNPs).

### Which continents exchanged SARS-CoV-2 with Europe and France?

Through several dozen distinct, dated and ancestrally reconstructed phylogenetic trees, we first studied the worldwide origins of SARS-CoV-2 transmissions into France and Europe for each of the time periods studied (**Figure 3**). Intra-continent and intra-France transmissions accounted for most of the total transmission events, representing 79.3% and 87.4% during the first and second half of 2020, respectively (**Figure 3—figure supplement 1**). From March 2020, the number of intra-territory transmissions lowly increased first in Asia before an acceleration of virus circulation in May. The first exponential increase of SARS-CoV-2 transmissions was observed in some European countries in early-March, then concomitantly in France and North America two weeks later. Intra-Europe and intra-France transmission levels were then very low between July and end-September until the beginning of the second European wave from October to December 2020. The rate of intra-North America transmission events remained very high from March to December. Intra-territory transmissions in South America started to increase from mid-April while those in Africa remained very limited. South America and Asia had a similar profile of intra-territory transmissions, as occurring at high levels between July and October followed by a slackening until December (**Figure 3A**). The profiles of intra-territory transmission events across the replicates and for each period investigated were positively correlated to the estimated number of deaths related to SARS-CoV-2 in each geographic region (Spearman’s rank correlation *p* < 0.001; r = 0.96 for the lowest correlation; **Figure 3—figure supplement 1**), thus confirming that our methodology adequately reflects spatial diffusion of COVID-19.

**Figure 3.**
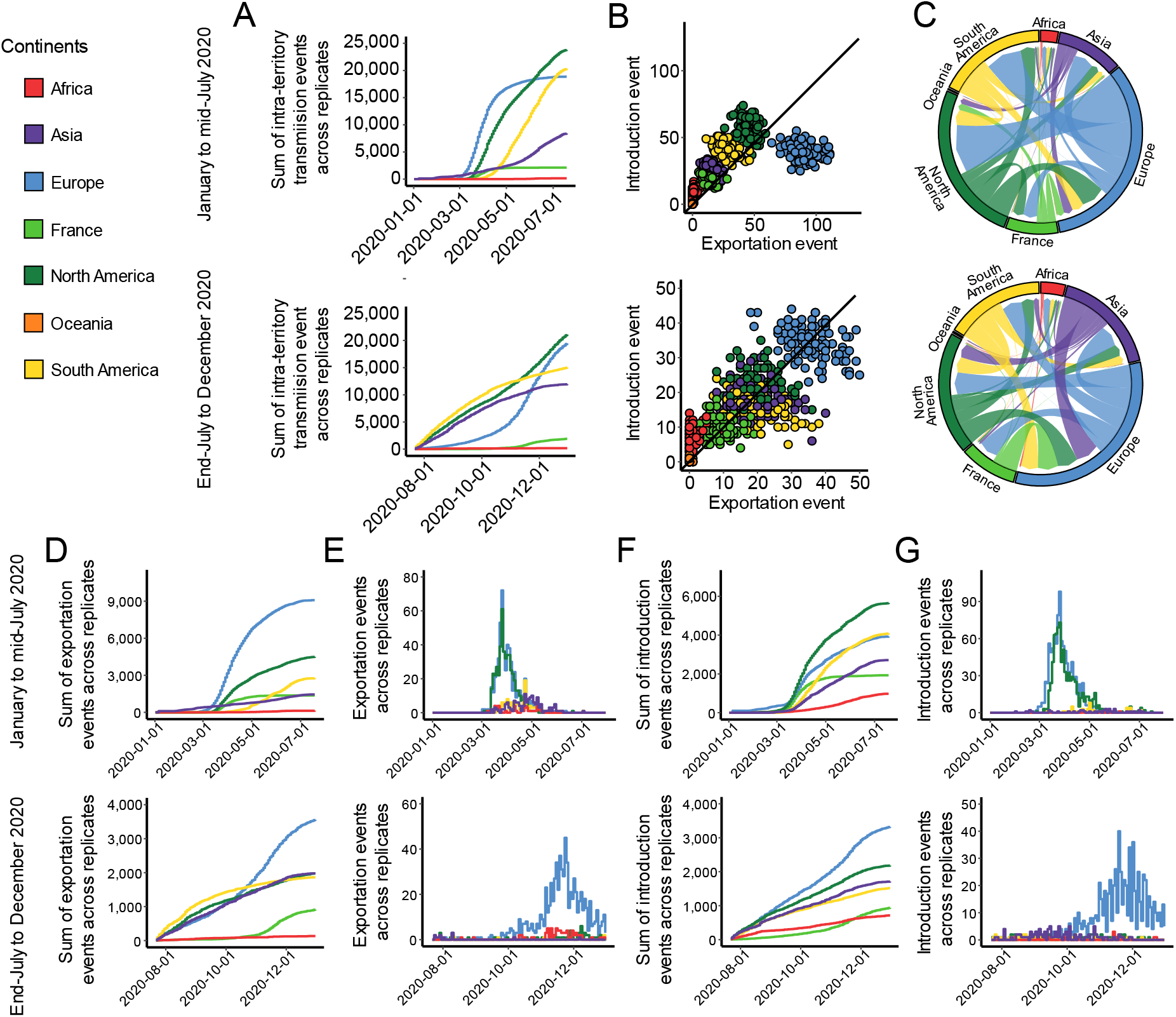
SARS-CoV-2 intra- and inter-territory transmissions worldwide. Each territory is associated with a specific color. Transmission events were calculated through 100 phylogenies as replicates between January and mid-July 2020, and between end-July to December 2020. When unspecified, transmission values are based on the sum of the replicates. (**A**) Evolution of intra-territory transmissions over time. (**B**) Number of introduction and exportation events for each replicate and for each continent and France. (**C**) SARS-CoV-2 exchange flows between continents and France during the two time periods investigated. In these plots, migration flow out of a particular location starts close to the outer ring and ends with an arrowhead more distant from the destination location. Migration flow out correspond to the mean of the worldwide sets of sequences. (**D**) Cumulative exportation and (**F**) introduction events per territory over time. (**E**) Exportation events over time originated from France. (**G**) Introduction events into France over time.

When focusing on introduction and exportation events during the first wave, we found that Europe was associated with the highest number of exportation events in all locations (**Figure 3B** and **3C**, and **Figure 3—figure supplement 2**). Both North America and South America also highly participated in virus exportation, while France and Asia had lower but similar numbers of exportation events across the replicates by the end of July 2020 (**Figure 3D** and **Figure 3—figure supplement 2**). The exportations from France were almost all headed towards Europe and North America (**Figure 3E** and **Figure 3—figure supplement 2**). North America accounted for a large proportion of SARS-CoV-2 introductions, followed by South America and Europe at similar rates by the end of July 2020 (**Figure 3F**). The first introduction cases into France were sparse and originated from Asia before March 2020, then introductions came mostly from Europe followed by North America and increased in number to reach a peak in end-March (**Figure 3G**). We noticed that the number of new introductions was low after mid-May 2020 in France until the second European epidemic wave.

The second half of 2020 (from end-July to December 2020) showed a very different pattern of inter-territory transmission events worldwide. The rate of exportation events was quite similar between North America, South America, Asia and Europe until mid-October, then this rate drastically increased only in Europe until the end of the time period investigated (**Figure 3B, 3C** and **3D**, and **Figure 3—figure supplement 2**). Before October 2020, rare exportation events originated from France, consistent with the very low incidence of the virus during this period in this country. Then, the number of virus exportations drastically increased and formed a peak in the mid of November 2020. Those exportations were nearly all headed towards Europe, accompanied with rare events into Africa and North America (**Figure 3E**). In the same way, only rare introduction events into France were originated from Asia, North America and South America until end-October, while high rates of introductions came from Europe from early-October to December 2020 (**Figure 3G**). SARS-CoV-2 introductions in Europe originated from Asia, North America, South America and France at similar levels (**Figure 3—figure supplement 2**).

### How did the virus spread in Europe?

We then aimed to get a finer view of SARS-CoV-2 transmission events between France and other European countries with the same strategy as the one used at the continental scale (**Figure 4**). Here, we only focused on European countries associated with a high incidence and without under-sampling due to a lack of data on GISAID.

**Figure 4.**
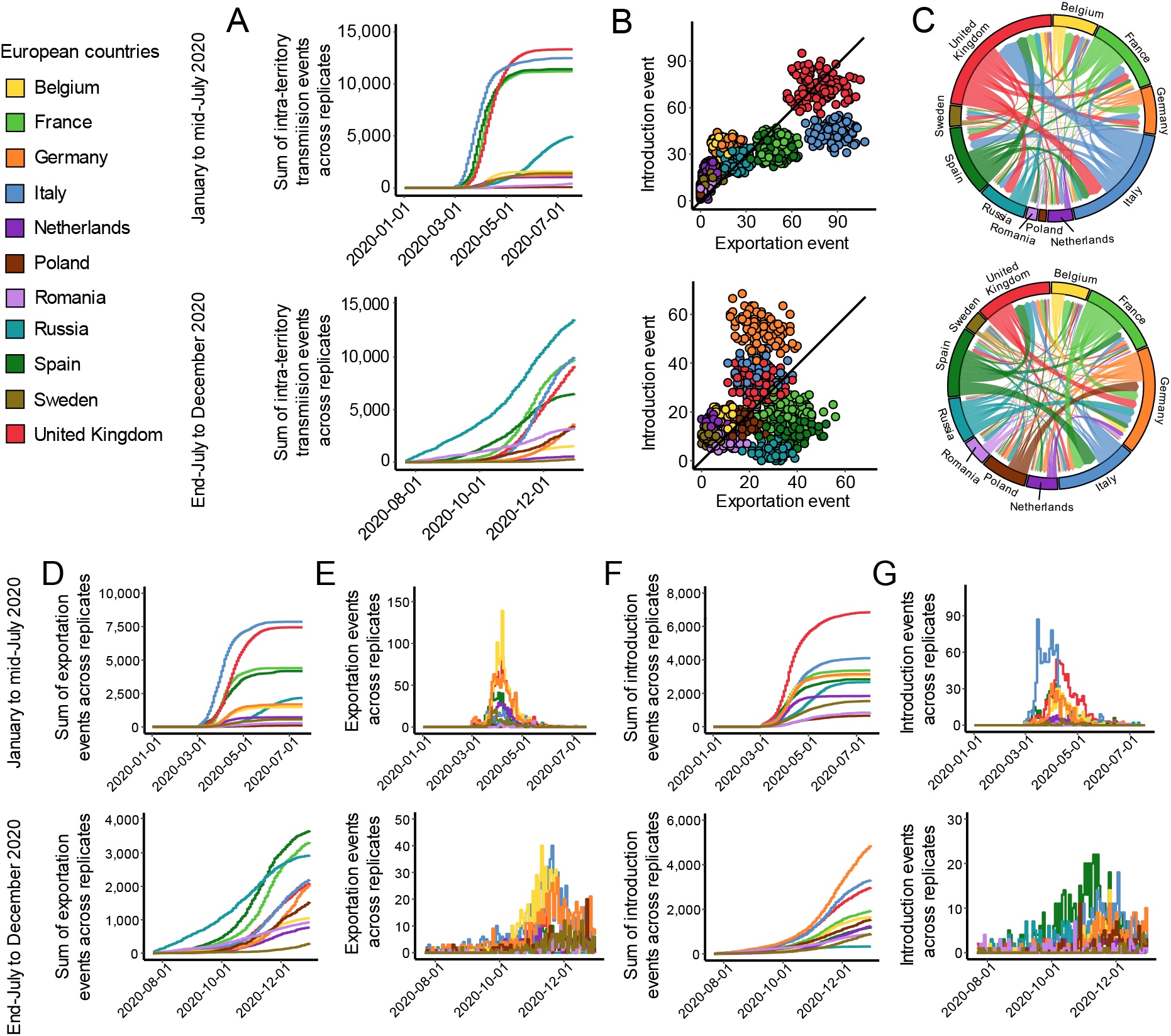
SARS-CoV-2 intra- and inter-territory transmissions at the European scale. Each territory is associated with a specific color. Transmission events were calculated through 100 phylogenies as replicates between January and mid-July 2020, and between end-July to December 2020. When unspecified, transmission values are based on the sum of the replicates. (**A**) Evolution of intra-territory transmissions over time. (**B**) Number of introduction and exportation events for each replicate and for each European country. (**C**) SARS-CoV-2 exchange flows between European countries during the two time periods investigated. In these plots, migration flow out of a particular location starts close to the outer ring and ends with an arrowhead more distant from the destination location. Migration flow out correspond to the mean of the sets of sequences. (**D**) Cumulative exportation and (**F**) introduction events per territory over time. (**E**) Exportation events over time originated from France. (**G**) Introduction events into France over time.

Intra-country transmissions accounted for most of the total transmission events, representing 65.7% and 75.6% of the transmissions during the first and second half of 2020, respectively (**Figure 4—figure supplement 1**). Between March and July 2020, the number of intra-country transmissions sequentially – and drastically – increased first in Italy, then Spain, France and the United Kingdom. Germany was associated with a number of intra-country transmissions as low as Belgium and the Netherlands (**Figure 4A** and **Figure 4—figure supplement 1**). While the number of intra-country transmission events reached a plateau in early-June 2020 across nearly all Europe, Russia and Romania were associated with an increased number of cases from April which coincided with a similar increase from Asian countries bordering Russia. The number of intra-country transmission events in Russia and Romania remained high until the end of December 2020. In early September 2020, Spain was the first country associated with a drastic increase of intra-territory transmissions, then followed by France, the United Kingdom, Italy and Poland. The increase was observed a bit later for Germany and Belgium, around end-October 2020. By the end of 2020, Russia accounted for most of the intra-country transmissions between July and December, followed by Italy and France, the United Kingdom and Spain. The profiles of intra-country transmissions across the replicates and for each wave investigated were, as for the continents, positively correlated to the estimated number of deaths related to SARS-CoV-2 for each country (Spearman’s rank correlation *p* < 0.001; r = 0.98 for the lowest correlation; **Figure 4—figure supplement 1**).

By calculating the count of introduction and exportation events during the first epidemic wave, we observed that Italy and the United Kingdom were the major contributor to virus dissemination towards other European countries, followed by France and Spain (**Figure 4B, 4C** and **4D**, and **Figure 4—figure supplement 2**). Italy was the first country to be associated with a drastic increase of exportation events in early-March (**Figure 4D**), followed by Spain, the United Kingdom and France about one week later. France and Spain were however surpassed by the United Kingdom by the end of April 2020. Most of exportation events from France were directed towards Belgium, the United Kingdom, and Germany, and a little towards Spain, the Netherlands and other European countries (**Figure 4C** and **Figure 4—figure supplement 2**), mostly before the official lockdown in France (i.e., before end-March) (**Figure 4E**). The rate of exportations from France then diminished until the second epidemic wave, as well as for other European countries except Russia. For introduction events, the United Kingdom accounted for a quarter of the total number of events, followed by Italy, while France, Germany, Spain and Belgium shared a strikingly similar pattern of transmissions (**Figure 4F**). In France, most of the introductions first came from Italy in early-March, while a few early introductions into France also originated from Spain, Germany and United Kingdom (**Figure 4G**). The United Kingdom, Spain, Belgium and Germany were associated with a high number of introductions into France, but mostly a few days before and at the beginning of the French national lockdown.

The second European epidemic wave showed a different pattern of transmission events. France, Spain and Russia were the countries associated with most of the exportation events, followed by Germany, Italy and The United Kingdom (**Figure 4B, 4C** and **4D**, and **Figure 4— figure supplement 2**). The profile of exportation events was globally uniform in Russia with a continuous increase through the whole year 2020 (**Figure 4D**). This was also the case in Romania, although the incidence remained lower than Russia. Spain, shortly followed by France, were the two first countries associated with a steep increase of exportations from September to December 2020. Later in September and October, the United Kingdom, Italy, followed by Poland, Germany, Belgium, and the Netherlands have each seen a sharp increase in their rate of exportations, sustained until the end of 2020. France moderately transmitted the virus until the end of October in some European countries, then a high rate of transmissions was mainly directed towards Belgium, Italy and Germany during November 2020 (**Figure 4E**). Concerning introduction events, Germany accounted for a large proportion of SARS-CoV-2 transmissions, followed by Italy and the United Kingdom (**Figure 4F** and **Figure 4—figure supplement 2**). For France, a few introductions came from Russia and Romania between August and September, while most of these events originated from Spain, Italy, Germany Belgium and the United Kingdom during November and December 2020 (**Figure 4G**).

Altogether, these results highlight that most governments adopted variable measures to contain COVID-19 pandemic between the first and second European epidemic waves with varying success rates. French people have, in all likelihood, traveled less abroad in Europe during the second wave.

### The first and second French epidemic waves: two patterns of SARS-CoV-2 transmissions within France

We finally conducted an in-depth analysis of virus spread inside France by studying transmission events between French administrative regions (**Figure 5**). As previously explained, only three regions (Auvergne-Rhône-Alpes, ARA, Île-de-France, IDF, and Provence-Alpes-Côte d’zur PACA, which cover the most densely populated cities in France, i.e., Paris, Lyon and Marseille) were investigated since only a few or in some cases no sequence at all were available on GISAID for the other regions (**Figure 2—figure supplement 1**).

**Figure 5.**
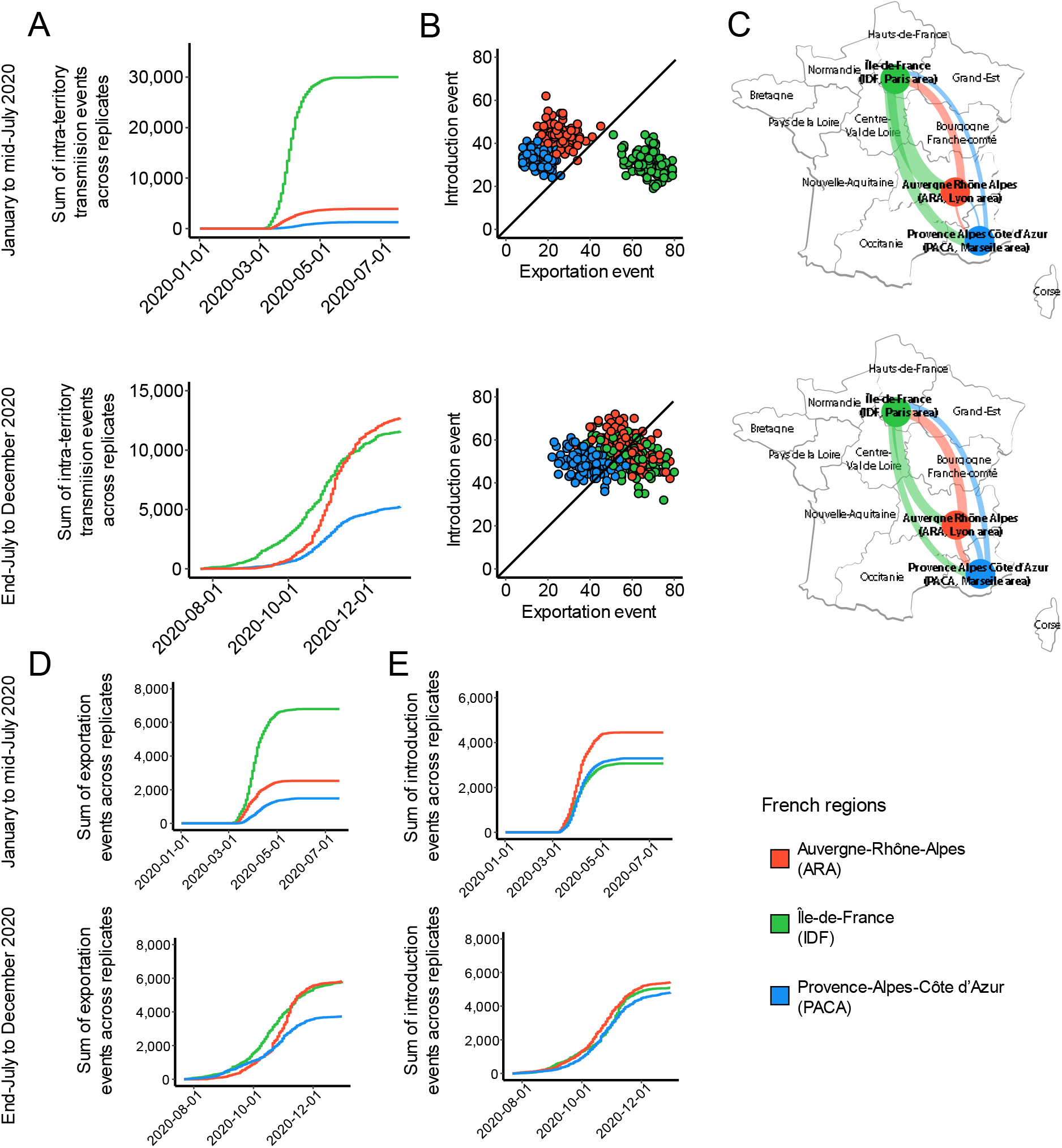
SARS-CoV-2 intra- and inter-territory transmissions in France. Each French administrative region is associated with a specific color. Transmission events were calculated through 100 phylogenies as replicates between January and mid-July 2020, and between end-July to December 2020. When unspecified, transmission values are based on the sum of the replicates. (**A**) Evolution of intra-region transmissions over time. (**B**) Number of introduction and exportation events for each replicate and for each French administrative region. (**C**) SARS-CoV-2 exchange flows between French administrative regions during the two time periods investigated. In these plots, migration flow out of a particular location starts close to the outer ring and ends with an arrowhead more distant from the destination location. Migration flow out correspond to the mean of the France sets of sequences. (**D**) Cumulative exportation and (**E**) introduction events per French administrative region over time.

Similarly to previous scales investigated, intra-region transmissions represented 75.5% of the transmissions during the first wave, but decreased to 66.3% during the second half of 2020 likely because of a much less strict national lockdown implemented by the end of the year compared to the first wave (**Figure 5—figure supplement 1**). IDF (Paris area) was associated with much more intra-region transmissions than ARA (Lyon area) and PACA (Marseille area) (**Figure 5A**). Both ARA and PACA had lower intra-than inter-region transmission events during the first wave. During this period, we counted seven and twenty-three times less transmission events in ARA and PACA than in IDF respectively, which was globally consistent with the reported count of SARS-CoV-2-related deaths reported in French hospitals within these regions (Spearman’s rank correlation *p* < 0.001; r = 0.97 for the lowest correlation; **Figure 5— figure supplement 1**). Furthermore, we observed that the first intra-region transmission events were found in IDF in early-March, then in ARA and PACA about one week later. The number of intra-region transmissions started to decrease from the end of March, fitting with the complete lockdown across the entire country that was enforced the 17th March 2020. Transmissions were very low during the summer 2020. Then, our results indicated a rise of intra-region transmissions from early September 2020 first in IDF, and three weeks later in ARA and PACA. During October, intra-region transmissions in ARA were higher than in IDF and PACA, an observation that contrasts with the first half of 2020. Intra-region transmissions in PACA were a bit lower than in IDF (**Figure 5—figure supplement 1**).

We then focused on introduction and exportation events during the first half of 2020. We observed three and four times more exportation events from IDF (Paris area) compared to ARA (Lyon area) and PACA (Marseille area), respectively (**Figure 5B** and **Figure 5—figure supplement 2**). Exportation events from IDF were a bit higher in ARA than in PACA (**Figure 5C**). ARA and PACA had more introduction events compared to IDF, while exportation events from ARA and PACA were mostly oriented towards IDF (**Figure 5C**). The first exportation events originated from IDF in early March, quickly followed, but at much lower rates, by ARA and a few days later by PACA (**Figure 5D**), while the first introduction events were observed in all locations (**Figure 5E**). Viral circulation started to decrease after the implementation of the national lockdown (17th March 2020). During the second half of 2020, IDF was no longer the single epicenter of SARS-CoV-2 inter-region transmissions. Both ARA and PACA participated concomitantly with IDF in virus spread at similar levels (**Figure 5B** to **5E**). Rare exportation and introduction events were detected in early August in all regions. The rate of exportation events rapidly increased first in IDF and PACA from mid-September, then in ARA from end-September and even exceeded other regions by the end of the year (**Figure 5D** and **5E**).

Altogether, SARS-CoV-2 transmission events within France showed two distinct patterns between the first and second half of 2020. While most of transmissions and subsequent deaths related to SARS-CoV-2 were localized in the vicinity of Paris during the first half of 2020, those indicators were more balanced after the summer 2020 with an important part of transmissions and deaths around Lyon (ARA) and Marseille (PACA) regions. This may be attributable to French people limiting journeys across Europe during the summer vacations to the benefit of enjoying the French landscapes.

## Discussion

Until now, the dynamics of SARS-CoV-2 transmissions within France or between France and other countries (in Europe or worldwide) remain only partially characterized. To our knowledge, three studies have explored the virus spread in France, but only focused on the first French epidemic wave and/or disregarded transmission events SARS-CoV-2 introductions from other continents or other European countries (Danesh et al., 2020; Elie and Alizon, 2020; Gámbaro et al., 2020). Other studies have focused on larger scale such as the whole Europe, but did not deeply explored the dynamics especially in France (Lemey et al., 2021; Nadeau et al., 2021). Here, we studied introduction and exportation events in France at different scales during the two first European epidemic waves. We think our approach provides a way to tackle large genomic data for future epidemics and allows a deeper comprehension of viral circulation in France and Europe.

Methodologically, most phylodynamics studies on SARS-CoV-2, as well as on other viruses or pathogens, are based on Bayesian approaches (notably with the popular and robust BEAST tool; Drummond and Rambaut, 2007) using a unique phylogenetic tree composed of several thousand sequences. To reconstruct the evolution parameters and ancestral states at nodes (here, the geographical area), Bayesian methods require high computational power, representing their main limitation (Drummond and Rambaut, 2007). Overall, the generation of a phylogenetic tree and the ancestral state reconstruction for even a few thousands of sequences is no small feat because requiring very long timeframe, or the parameter estimation can fail to converge. Furthermore, conclusions obtained will be based on a single sampling with a relatively limited number of sequences, but different samplings may lead to some contradictory results (Hall et al., 2016). Since the GISAID database stores several hundred thousand SARS-CoV-2 sequences (n = 638,704 on the 8th May 2020 for the year 2020 alone), we believe that one phylogeny of a few thousand sequences only may not always accurately reflect transmission events. In order to obtain more comprehensive trends drawing conclusions from a maximum of SARS-CoV-2 sequences, we constructed 100 independent maximum likelihood phylogenies of relatively limited size (from 462 to 999 sequences depending on the geographic scale and time period investigated) and inferred ancestral state reconstruction by a maximum likelihood discrete trait phylogeographic method. Bayesian approach seems not to be more accurate than maximum likelihood for the reconstruction of ancestral states (Hanson-Smith et al., 2010), and we gain a lot in terms of sampling, computational time, and trends through distinct tens of replicates. As a direct validation of our approach, we observed that the number of intra-transmission events (i.e., a SARS-CoV-2 transmission within a same territory) was directly correlated to the number of SARS-CoV-2-related deaths reported by national health agencies. Also, the method was recently used to depict with accuracy the genomic epidemiology of the virus in Canada (McLaughlin et al., 2022).

The pipeline conducted here allowed to get an in-depth view of SARS-CoV-2 transmissions implying France at both local and international contexts during 2020. During the first European epidemic wave, France mostly exchanged SARS-CoV-2 with Europe and North America. The observation contrasts with the second European epidemic where France exclusively shared the virus with Europe. This result is quite logical insofar as the borders of France have gradually reopened with European countries, but not with the rest of the world. A very low rate of SARS-CoV-2 transmissions between Europe (including France) and Asia or South America was found in 2020, which is not surprising since the virus started to worryingly spread in these continents only from May 2020 where all borders of Europe were closed. At the European scale, France was unambiguously a major actor of SARS-CoV-2 transmission. It was ranked third regarding exportation events (mainly directed towards neighboring countries) only behind the United Kingdom and Italy during the first wave, and second behind Spain during the second one. Temporally, the first introduced European cases into France came from Italy and Spain, consistent with the beginning of SARS-CoV-2 spread in Europe. These results are globally in line with the observations made across Europe through a Bayesian inference (Lemey et al., 2021). For the second European wave, a few introduced cases into France came from Russia (and at lower rates, from Romania) which was associated with an impressive increase in virus incidence since the summer 2020. Then, most virus introductions into France originated from Spain which initiated the second wave in European Union, followed by Italy and Germany. Interestingly, a much higher number of intra-country transmission events was systematically noticed during the second wave compared to the first one, indicating the effectiveness of border control measures and that French people globally not travelled outside France after the first national lockdown.

Our results at the French scale should be interpreted in light of several limitations. In addition to a limited overall size, genomic data only cover three out of thirteen administrative regions (**Figure 2—figure supplement 1**). The North and East of France (i.e., regions Hauts-de-France and Grand-Est, respectively) that border Belgium, Luxembourg and Germany, were not investigated. More importantly, the region Grand-Est was associated with early large clusters that participated in virus dissemination across the country during the first European epidemic wave, such as the evangelical gathering of the Christian Open-Door Church (Mulhouse, Haut-Rhin) that took place from February 17th to 21th 2020 and consisted of approximately a thousand contaminated believers. Keeping these limitations in mind, we observed two distinct patterns of SARS-CoV-2 transmissions within France during 2020. During the first European wave, most of the transmissions originated from Île-de-France (IDF; Paris area). Many people living in IDF decided to leave the Paris region at the beginning of the epidemic to live in their second homes in the provinces, which seems to have resulted in a wide spread of the virus seeded from Paris. For the second epidemic wave, we observed a balanced transmission contribution between IDF and the other regions, that can be explained as most people from IDF still resided in the provinces without returning to IDF during the second part of 2020.

Overall, using an original approach allowing deeply investigate SARS-CoV-2 phylodynamics taking into account a very large number of sequences, our findings allow a stronger understanding of SARS-CoV-2 transmission events related to France and Europe in 2020 and the changes observed between the two epidemic waves.

## Methods

### Ethics statement

This study was carried out in accordance with the Declaration of Helsinki. It is a retrospective non-interventional study based on data from the public database repository GISAID (https://www.gisaid.org/). The Institutional Review Board – IRB 00006477 – of HUPNVS, Université Paris Cité (AP-HP), has reviewed and approved the research project. Laboratories and organisms that participated in depositing SARS-CoV-2 sequences used in this study can be found on the GISAID database from the identifiers listed in **Supplementary Dataset 1**.

### Sequence acquisition and curation, and dataset production

A phylodynamics study (which aims to depict with accuracy transmission events between locations; **Figure 6**), requires a large number of sequences (Attwood et al., 2022). SARS-CoV-2 genome sequences were downloaded from the GISAID database on the 8th May 2022. The following inclusion and exclusion criteria were applied to select samples for analysis: (1) only complete viral genomes from infected individuals (more than 29 kb in length) were included; (2) the maximal proportion of undetermined nucleotide bases was fixed to 5%; (3) sequences with unknown or incomplete sampling dates, or unknow geographical location, were excluded; sampling dates were comprised between January 1st 2020 and December 31th 2020; and sequences with host listed as anything but were excluded. The final dataset was constituted of 638,704 sequences.

**Figure 6.**
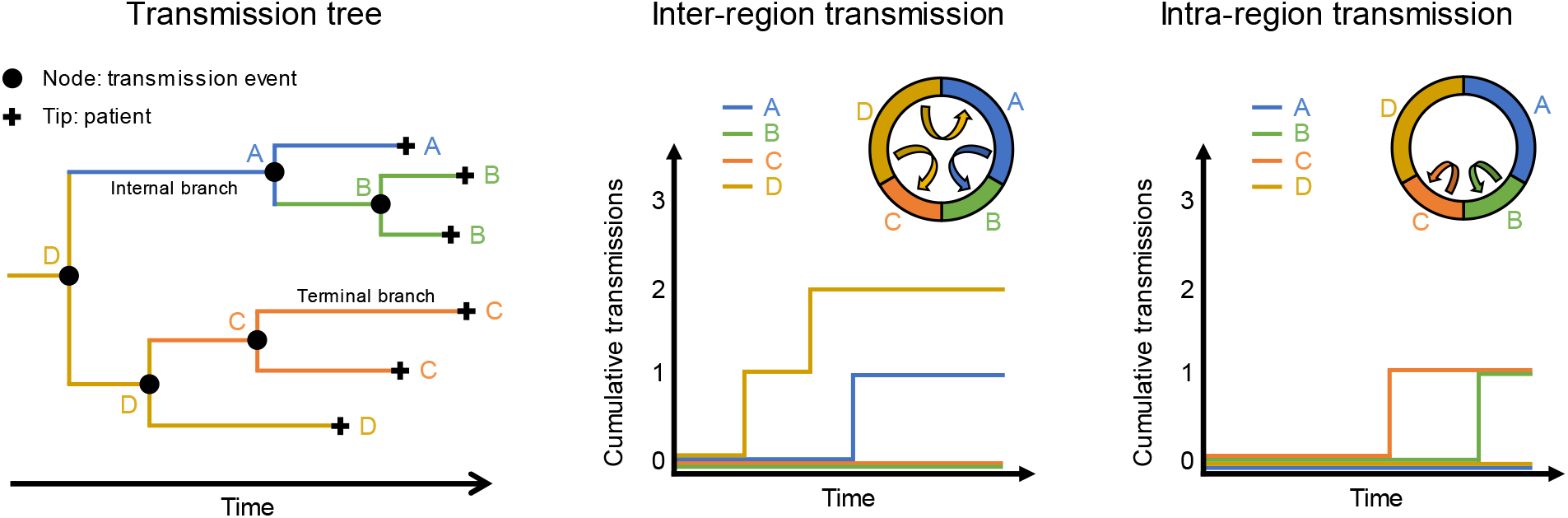
Conceptual overview of phylodynamic analysis. Each tip of a phylogenetic tree corresponds to a patient infected by SARS-CoV-2 in a given geographical location and at a given date. Through maximum likelihood, each node may be assigned with a geographical location and each branch from this node correspond to a transmission event within this location, if the same location is also observed at the next node, or, in the contrary, from the ancestral node location to the following node location. The sum of branch lengths to root may be used to calculate the date of transmission. The number of transmissions inside or outside a same geographical location could then be established.

We studied the epidemic at three different geographic scales: (1) worldwide, to identify SARS-CoV-2 transmissions from continents to France and conversely; (2) at the European scale, to determine more precisely which European countries were mostly involved in transmission flows with France; and finally (3) at the French administrative region level, to get a better understanding of virus spread inside the country. For each geographic scale, the analyses were independently run for the two European epidemic waves (January to mid-July 2020 (weeks 1 to 30), and end-July to December 2020 (weeks 31 to 52); **Figure 1**).

Considering the large number of sequences in GISAID that passed our criteria (n = 638,704), we produced for each investigated geographic scale analysis (i.e., worldwide, Europe, and French administrative regions) and time period (January to mid-July 2020 and end-July to December 2020) 100 distinct sets of sequences. To determine the number of SARS-CoV-2 genome sequences to be included so as to be representative of each location epidemic, we used the number of SARS-CoV-2-related deaths per territory and per week as a proxy for the number of infections taking place two weeks earlier. For each set, sequences were drawn randomly from our GISAID extraction without replacement. Some countries and French regions had to be discarded because of their under-representation in the GISAID database. When a set was constituted, all the sequences were replaced in the whole dataset for the random selection of the next one. For the first time period investigated (January to mid-July 2020), an average of 935, 999 and 462 SARS-CoV-2 sequences were included in each set at the worldwide, European and French scales, respectively (**Table 1**). When considering the second time period (end-July to December 2020), we used about 300 sequences from the first wave to anchor phylogenies with the previously acquired viral diversity. In addition to those sequences, we added worldwide, European and French scale sequence dataset containing 882, 961 and 483 genomic sequences covering July to December 2020 (**Table 1**).

### Multiple sequence alignment and maximum likelihood phylogenetic tree

For each set, genome sequences were each independently aligned against the Wuhan-Hu-1 reference genome (GenBank accession: NC_045512.2) using MAFFT v7.450 then merged altogether (Katoh and Standley, 2013). From each alignment, we inferred a maximum likelihood phylogenetic tree using FastTree v.2.1.11 under a general time-reversible (GTR) model with a discrete gamma distribution to model inter-site rate variation and 1,000 bootstraps to compute support values (Price et al., 2010). The choice of FastTree was mainly due to its short runtime while giving results very close to IQTree 2.1.2 (Minh et al., 2020). Each phylogenetic tree was dated using TreeTime v.0.8.5 (Sagulenko et al., 2018) with a molecular clock rate fixed at 6.00 × 10^−3^ substitutions per site per year. Note that this evolutionary rate is not in agreement with the one largely reported in other studies (clock rate of ∼ 6.00 × 10^−4^ substitutions per site per year; Xia, 2021), but it permits to date the tree without overestimation of branch lengths (**Figure 6—figure supplement 1**). Phylogenetic trees were manipulated and viewed using treeio and ggtree R packages respectively (Wang et al., 2020; Yu et al., 2017).

### Worldwide, Europe and French region SARS-CoV-2 phylodynamics analyses

For each phylogenetic tree we produced, ancestral state reconstructions of discrete, geographical data were performed with the *ace* function of ape package in R (Paradis et al., 2004). Three distinct Markov models of discrete character evolution through maximum likelihood were tested and compared: the Equal Rates (ER) which assumes a single rate of transition among all possible states, the All Rates Different (ARD) which allows a distinct rate for each position transition between two states, and the Symmetrical Rates (SYM) where forward and reverse transitions share the same parameter. In this study, we only present the results obtained with the SYM model since the ARD model led to high variations across replicates probably because the model is too sensible to outliers (**Figure 6—figure supplement 2**), while the ER model led to nearly identical observations compared to the SYM model (data not showed).

With the reconstruction, we assigned nodes to a location when supported ≥50% the state assignment. To detect and count each transition event, we checked for each node of the phylogenetic tree whether children nodes corresponded to the same geographic region as the current node, i.e., a SARS-CoV-2 intra-territory transmission (transmission event that does not cross any administrative border). If they correspond to different geographic locations, we then assumed the transmission from the parent node-state to the given child node-state (SARS-CoV-2 inter-territory transmission). The midpoint of the parent-child branch was chosen as the date of transmission in all cases (**Figure 6**).

## Data Availability

Data produced in the present work are contained in the manuscript. Large datasets are available upon reasonable request to the authors.

## Declaration

### Consent for publication

There are no case presentations that require disclosure of respondent‘s confiential data/information in this study.

### Availability of data and materials

All sequences used in the study are publicly available on the GISAID database. The list of the GISAID identifiers for each dataset is provided in the **Supplementary Data 1**. All the scripts developed for this study were deposited in the following GitHub repository: https://github.com/Rcoppee/PhyloCoV. Large metadata can be shared on demand.

### Author’s contributions

**Romain Coppée:** Conceptualization, Methodology, Software, Validation, Formal analysis, Investigation, Resources, Data curation, Writing – Original Draft, Writing – Review & Editing, Visualization. **François Blanquart:** Software, Data curation, Writing – Review & Editing. **Aude Jary:** Writing – Review & Editing. **Valentin Leducq:** Writing – Review & Editing. **Valentine Marie Ferré:** Writing – Review & Editing. **Anna Maria Franco Yusti:** Writing – Review & Editing. **Lena Daniel:** Writing – Review & Editing. **Charlotte Charpentier:** Writing – Review & Editing. **Samuel Lebourgeois:** Writing – Review & Editing. **Karen Zafilaza:** Writing – Review & Editing. **Vincent Calvez:** Writing – Review & Editing. **Diane Descamps:** Writing – Review & Editing. **Anne-Geneviève Marcellin:** Writing – Review & Editing. **Benoit Visseaux:** Conceptualization, Methodology, Validation, Writing – Review & Editing, Supervision, Project administration, Funding acquisition. **Antoine Bridier-Nahmias:** Conceptualization, Methodology, Software, Validation, Formal analysis, Investigation, Resources, Data curation, Writing – Review & Editing, Supervision, Project administration

## Acknowledgements

We gratefully thank all the laboratories and organisms that deposited SARS-CoV-2 sequences on the GISAID dataset, without which this study would not have been possible. All providers of sequences used in this study are available on GISAID from the accession numbers listed in the **Supplementary Data 1**.

## Figure supplement legends

**Figure 1—figure supplement 1**. SARS-CoV-2-related deaths per week of 2020 for each (**A**) continent and (**B**) European country. Each color corresponds to a territory.

**Figure 2—figure supplement 1**. Differential profiles of SARS-CoV-2 sequences wanted and available for each French region. In this analysis, we requested 500 SARS-CoV-2 for each time period (January to mid-July 2020 and end-July to December 2020) to generate a dataset covering the different French administrative regions. We calculated the differential between the number of sequences available and the number of sequences that are required to properly represent the number of SARS-CoV-2 infections per week. Green and red colors correspond to an excess and a lack of sequences for a week, respectively. For each region, we also calculated the percentage of weeks where a lack of sequences was observed. For each time period, we also indicated the number of sequences wanted.

**Figure 2—figure supplement 2**. Within-territory pairwise genetic distance across the replicates for each period at (**A**) worldwide, (**B**) European and (**C**) French administrative region scales. The first row of plots corresponds to the data from January to mid-July 2020, and the second one covers the period of end-July to December 2020. Each color corresponds to a territory. Distances were calculated from the dated trees (with TreeTime) using the *distTips* function of adephylo R package. Abbreviations: ARA, Auvergne-Rhône-Alpes; IDF, Île-de-France; PACA, Provence-Alpes-Côte d’zur

**Figure 3—figure supplement 1**. Intra-territory SARS-CoV-2 transmissions and correlation with epidemiological data at the worldwide scale. (**A**) SARS-CoV-2 inter- and intra-territory flows during the first and second half of 2020. In these plots, migration flow out of a particular location starts close to the outer ring and ends with an arrowhead more distant from the destination location. Migration flow out correspond to the mean of the worldwide sets of sequences. (**B**) Dynamics of intra-territory transmission events across the 100 independent replicates during the first (*left plot*) and second (*right plot*) half of 2020. Each territory is associated with a specific color. Profiles are very similar to the estimated numbers of SARS-CoV-2-related deaths. The curves were smoothed based on a window of seven days. (**C**) Correlation between intra-territory events and the number of SARS-CoV-2-related deaths over time. In a graph, a point corresponds to the cumulative numbers of intra-territory transmissions and the number of deaths at a given date. Each plot and corresponding color correspond to a territory. The coefficient of correlation R is indicated inside the plots.

**Figure 3—figure supplement 2**. Inter-territory SARS-CoV-2 transmissions at the worldwide scale. (**A**) Variations of exportation and introduction events across the 100 independent replicates. The two first plots show the variation of the events on the data covering January to mid-July 2020, while the two last plots display the variation between End-July and December 2020. Each color corresponds to a territory. The standard deviation corresponds to the sum of the previous replicates. The first replicates are associated with high variations, but tendencies are rapidly reached. (**B**) Mean number of introduction and exportation events for each continent and France. During the first half of 2020, Europe has much more exportation than introduction events. Between end-July and December 2020, all territories harbored nearly equal exportation and introduction events. (**C**) Number of exportation events from France and (**D**) introduction events into France according to the different continents. (**E**) Worldwide maps of continental and France transmissions. The width of the links is proportional to the number of transmission events. The color, specific to each territory, indicates the flow direction. The points that serve to anchor the territories were randomly placed.

**Figure 4—figure supplement 1**. Intra-territory SARS-CoV-2 transmissions and correlation with epidemiological data at the European scale. (**A**) SARS-CoV-2 inter- and intra-territory flows during the first and second half of 2020. In these plots, migration flow out of a particular location starts close to the outer ring and ends with an arrowhead more distant from the destination location. Migration flow out correspond to the mean of the sets of sequences. (**B**) Dynamics of intra-territory transmission events across the 100 independent replicates during the first (*left plot*) and second (*right plot*) half of 2020. Each European country is associated with a specific color. Profiles are very similar to the estimated numbers of SARS-CoV-2-related deaths. The curves were smoothed based on a window of seven days. (**C**) Correlation between intra-territory events and the number of SARS-CoV-2-related deaths over time. In a graph, a point corresponds to the cumulative numbers of intra-territory transmissions and the number of deaths at a given date. Each plot and corresponding color correspond to an European country. The coefficient of correlation R is indicated inside the plots.

**Figure 4—figure supplement 2**. Inter-territory SARS-CoV-2 transmissions at the European scale. (**A**) Variations of exportation and introduction events across the 100 independent replicates. The two first plots show the variation of the events on the data covering January to mid-July 2020, while the two last plots display the variation between End-July and December 2020. Each color corresponds to an European country. The standard deviation corresponds to the sum of the previous replicates. The first replicates are associated with high variations, but tendencies are rapidly reached. (**B**) Mean number of introduction and exportation events for each European country. (**C**) Number of exportation events from France and (**D**) introduction events into France according to the different European countries. (**E**) Maps of European SARS-CoV-2 transmissions. The width of the links is proportional to the number of transmission events. The color, specific to each European country, indicates the flow direction. The points that serve to anchor the territories were randomly placed.

**Figure 5—figure supplement 1**. Intra-territory SARS-CoV-2 transmissions and correlation with epidemiological data at the France scale. (**A**) SARS-CoV-2 inter- and intra-territory flows during the first and second half of 2020. In these plots, migration flow out of a particular location starts close to the outer ring and ends with an arrowhead more distant from the destination location. Migration flow out correspond to the mean of the worldwide sets of sequences. (**B**) Dynamics of intra-territory transmission events across the 100 independent replicates during the first (*left plot*) and second (*right plot*) half of 2020. Each territory is associated with a specific color. Profiles are very similar to the estimated numbers of SARS-CoV-2-related deaths. The curves were smoothed based on a window of seven days. (**C**) Correlation between intra-territory events and the number of SARS-CoV-2-related deaths over time. In a graph, a point corresponds to the cumulative numbers of intra-territory transmissions and the number of deaths at a given date. Each plot and corresponding color correspond to a territory. The coefficient of correlation R is indicated inside the plots.

**Figure 5—figure supplement 2**. Inter-territory SARS-CoV-2 transmissions at the France scale. (**A**) Variations of exportation and introduction events across the 100 independent replicates. The two first plots show the variation of the events on the data covering January to mid-July 2020, while the two last plots display the variation between End-July and December 2020. Each color corresponds to a territory. The standard deviation corresponds to the sum of the previous replicates. The first replicates are associated with high variations, but tendencies are rapidly reached. (**B**) Mean number of introduction and exportation events for each French administrative region. During the first half of 2020, IDF has much more exportation than introduction events. Between end-July and December 2020, all territories harbored nearly equal exportation and introduction events.

**Figure 6—figure supplement 1**. Fixing branch lengths using a fixed clock-rate. We used the worldwide dataset covering the whole year 2020, and we focused on the first replicate as an example. (**A**) Maximum likelihood phylogenetic trees obtained after dating the trees using TreeTime, with estimated (*left*) and fixed (*right*) clock-rates. Colors at nodes correspond to dates (gradient color). (**B**) Date of tips and nodes retrieved from the phylogenies after applying an estimated (*left*) or a fixed (*right*) clock-rate. With the estimated clock-rate, consistent with the known evolutionary rate of SARS-CoV-2 (here, 5.80 × 10^−4^ substitutions per site per year), most nodes are dated between February and April 2020. Very rare nodes were dated after October 2020, despite that some samples were collected after this date. Most samples had very long branches. With the fixed clock-rate (here, 6.00 × 10^−3^ substitutions per site per year), branch lengths were much short and node dating was consistent with the date of sample collection.

**Figure 6—figure supplement 2**. High variations in the number transmission events using the ARD model. We used the worldwide dataset covering the whole year 2020 as an example. (**A**) Variations of exportation and introduction events across the 100 independent replicates. Each color corresponds to a territory. The standard deviation corresponds to the sum of the previous replicates. (**B**) Number of introduction and exportation events for each replicate and for each continent and France. The number of exportation events is very dispersal in the ARD model compared to both SYM and ER models, especially for Africa and Oceania. (**C**) Cumulative exportation events per territory over time. Across the replicates, Africa had similar exportation events than South America. This result is very unlikely since the virus much more circulated in South America compared to Africa. Another unlikely observation is that France had only two times less exportation events than Europe, suggesting that the single France contributed to the half of the exportation events.

