## Supplementary figures and images for "Phylodynamics of SARS-CoV-2 transmissions in France, Europe and the world during 2020"

### figure_1-supplement_fig_1.pdf

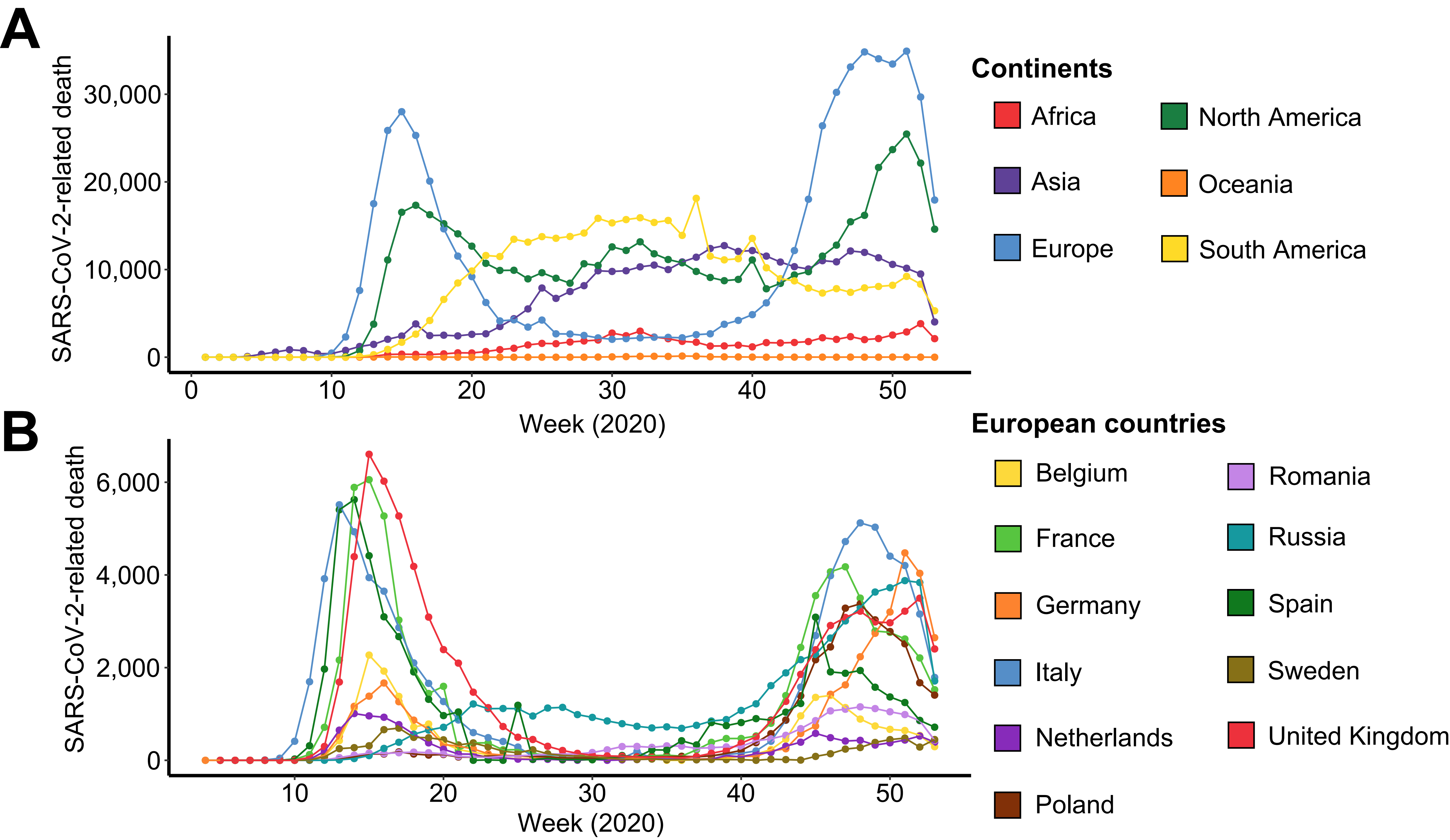

### figure_2-supplement_fig_1.pdf

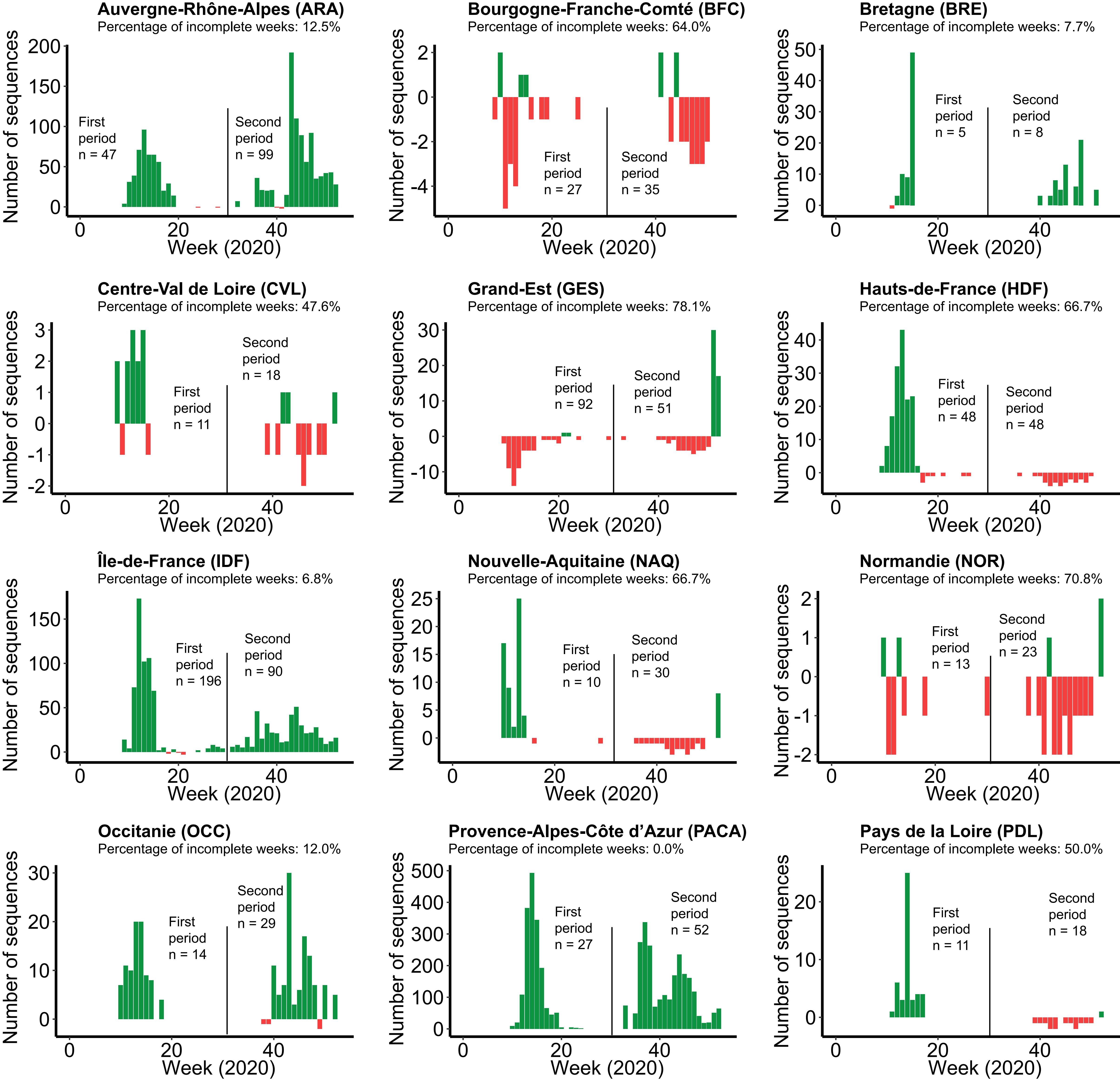

### figure_2-supplement_fig_2.pdf

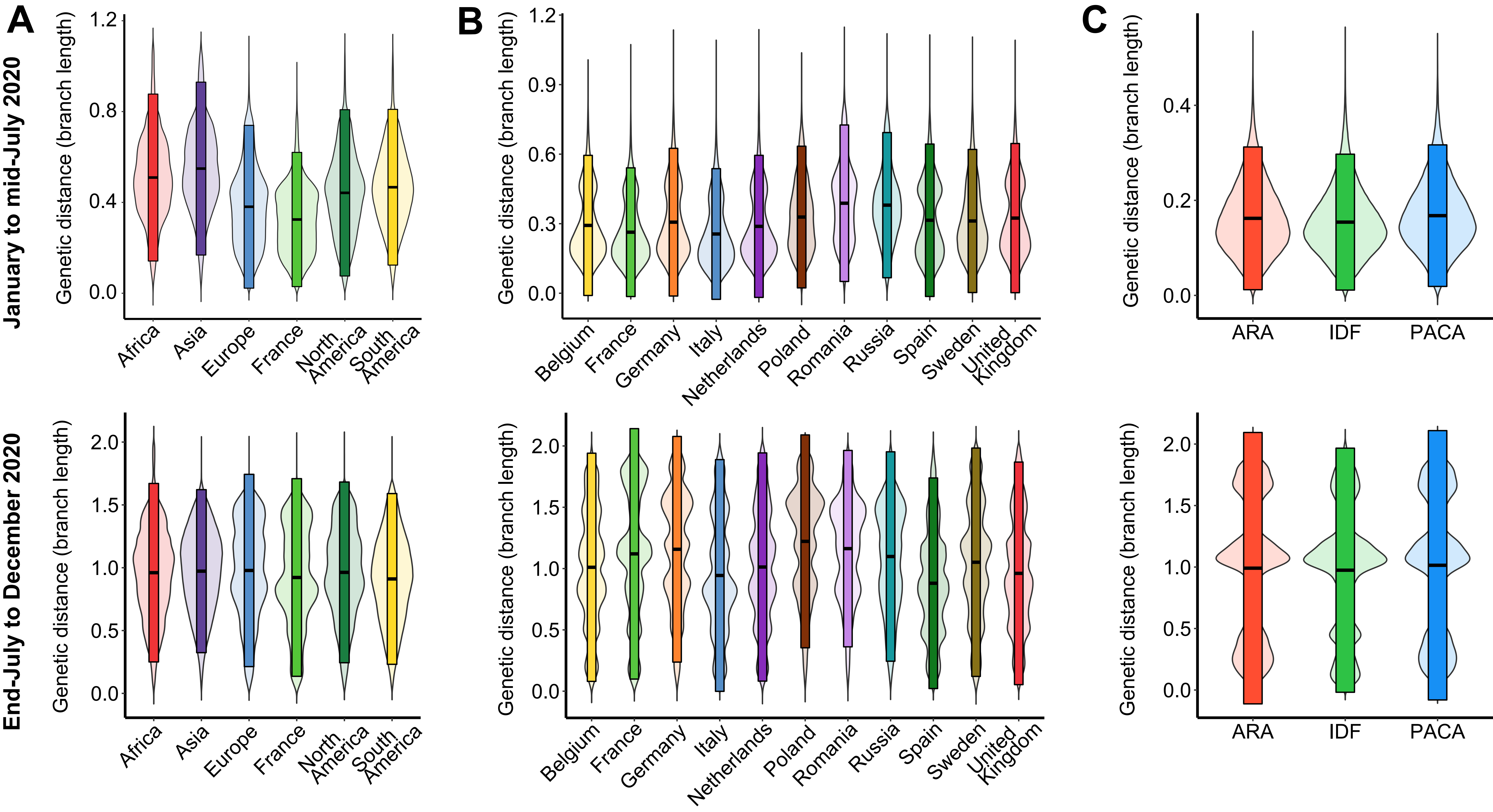

### figure_3-supplement_fig_1.pdf

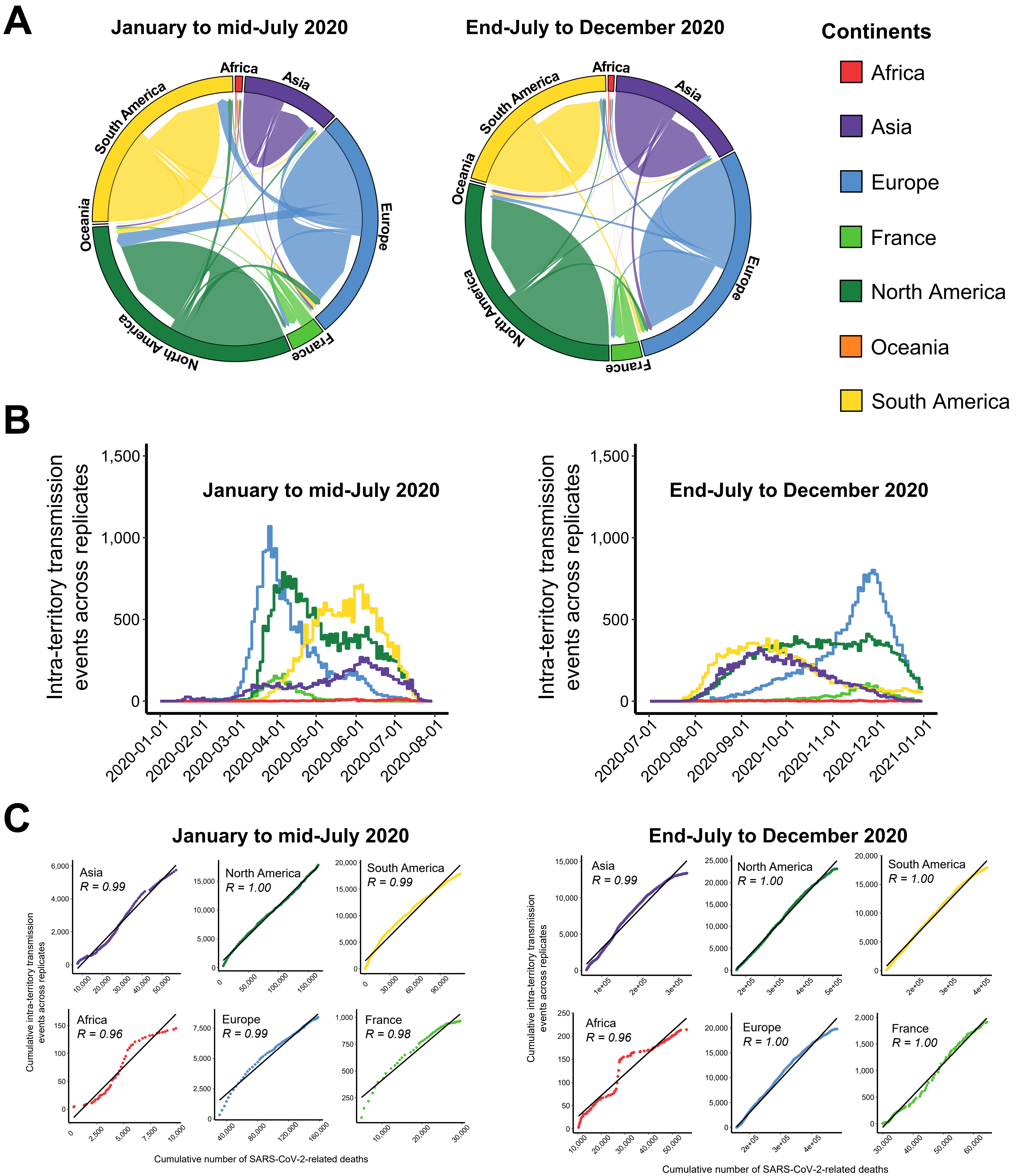

### figure_3-supplement_fig_2.pdf

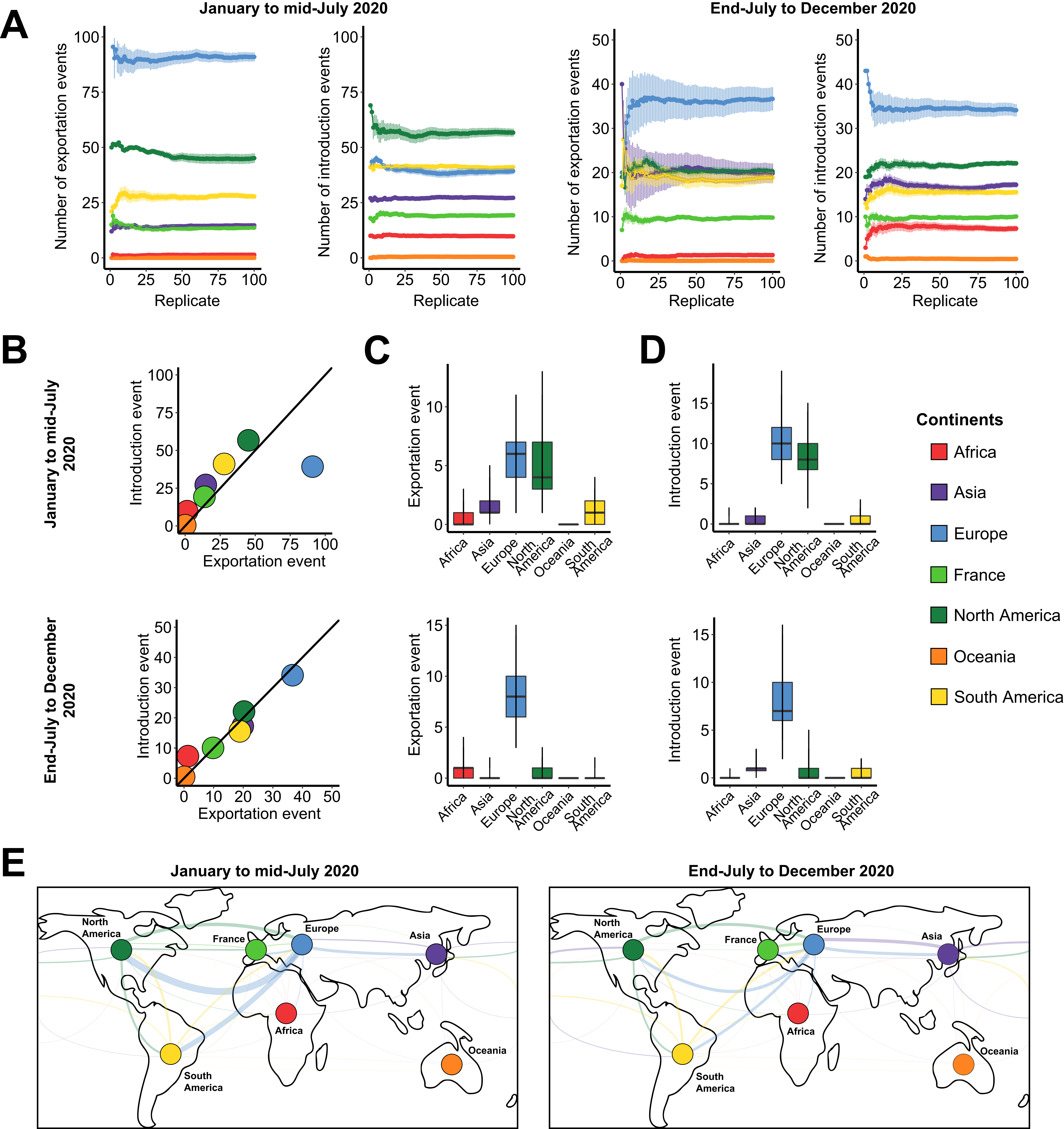

### figure_4-supplement_fig_1.pdf

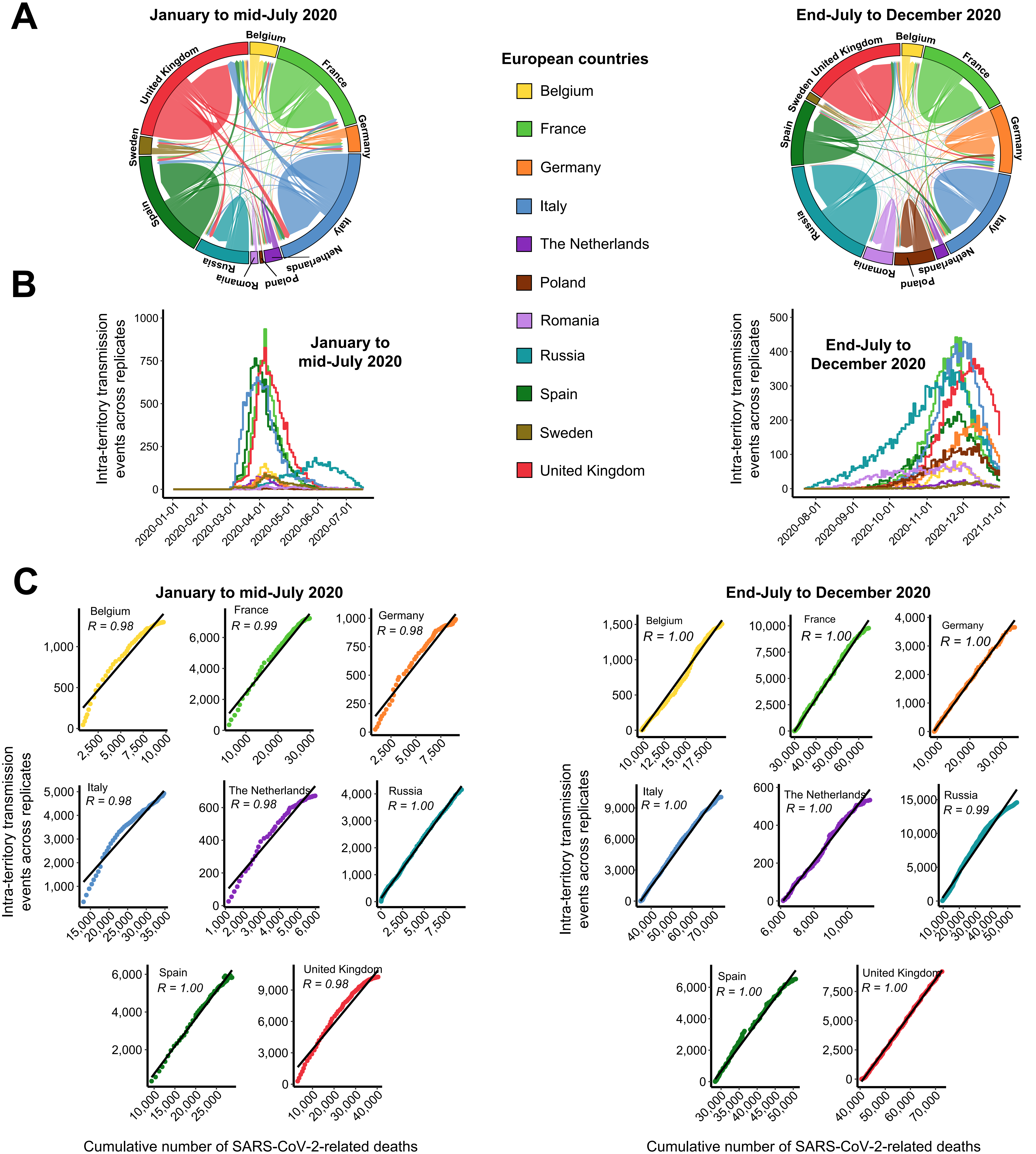

### figure_4-supplement_fig_2.pdf

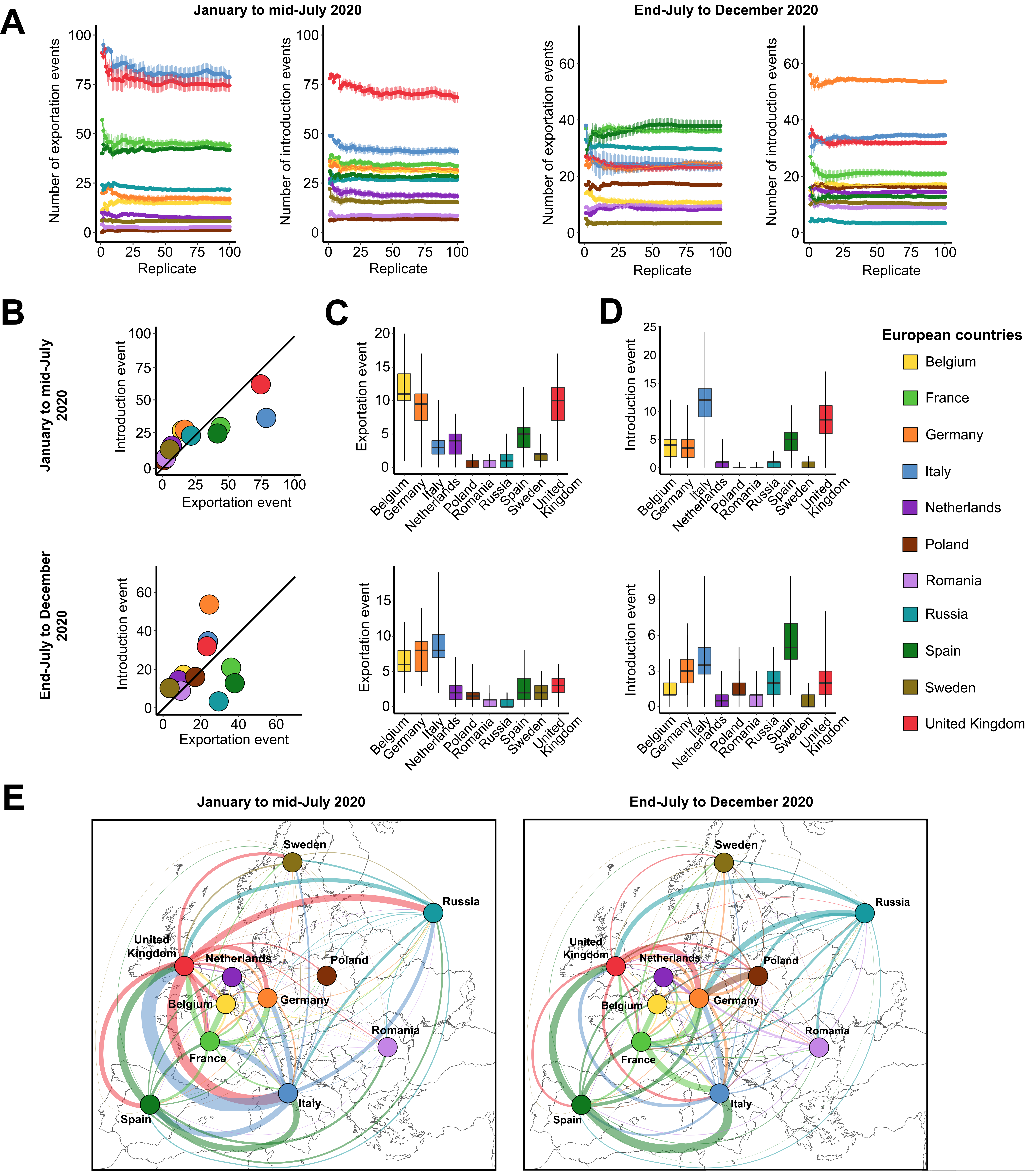

### figure_5-supplement_fig_1.pdf

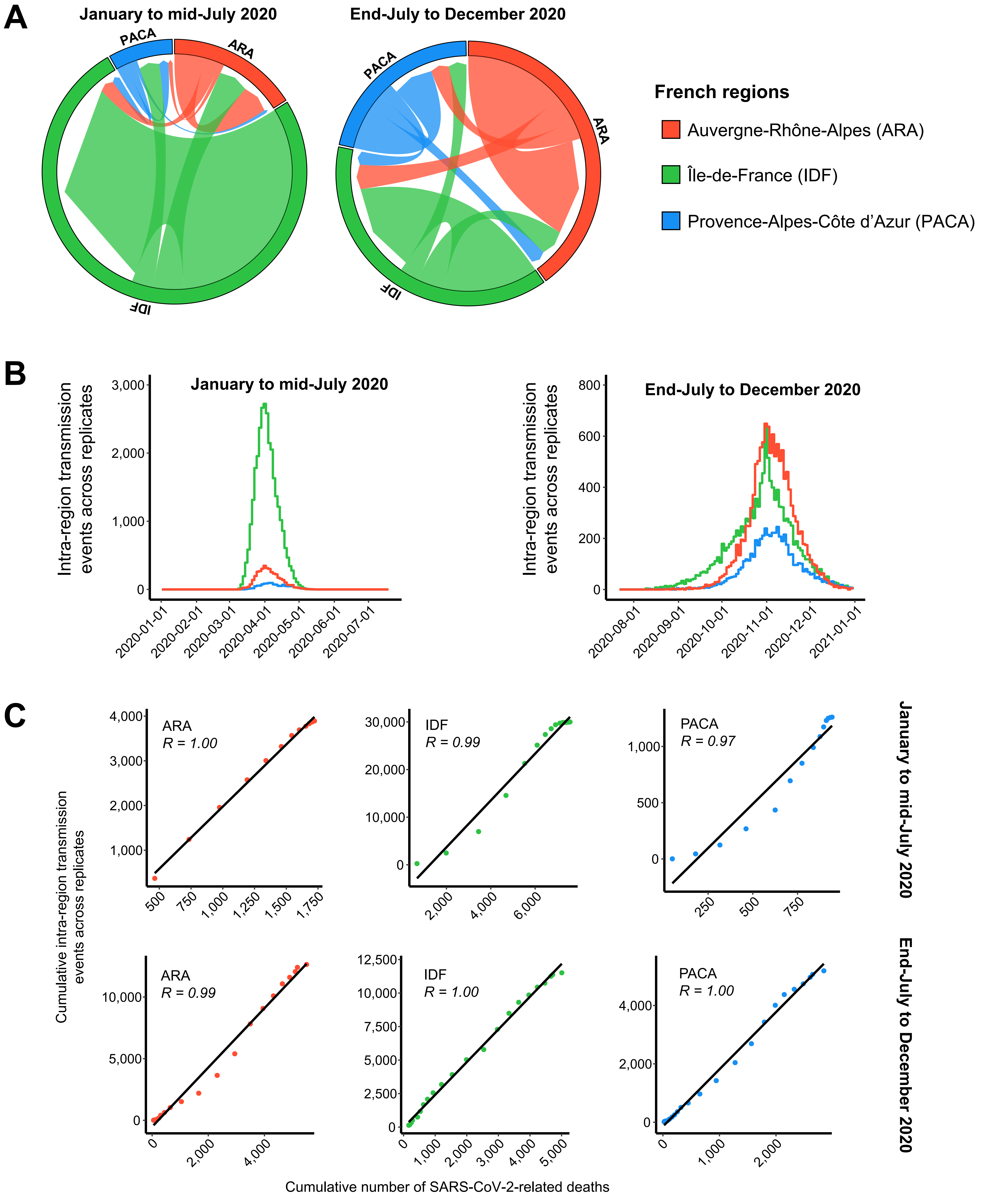

### figure_5-supplement_fig_2.pdf

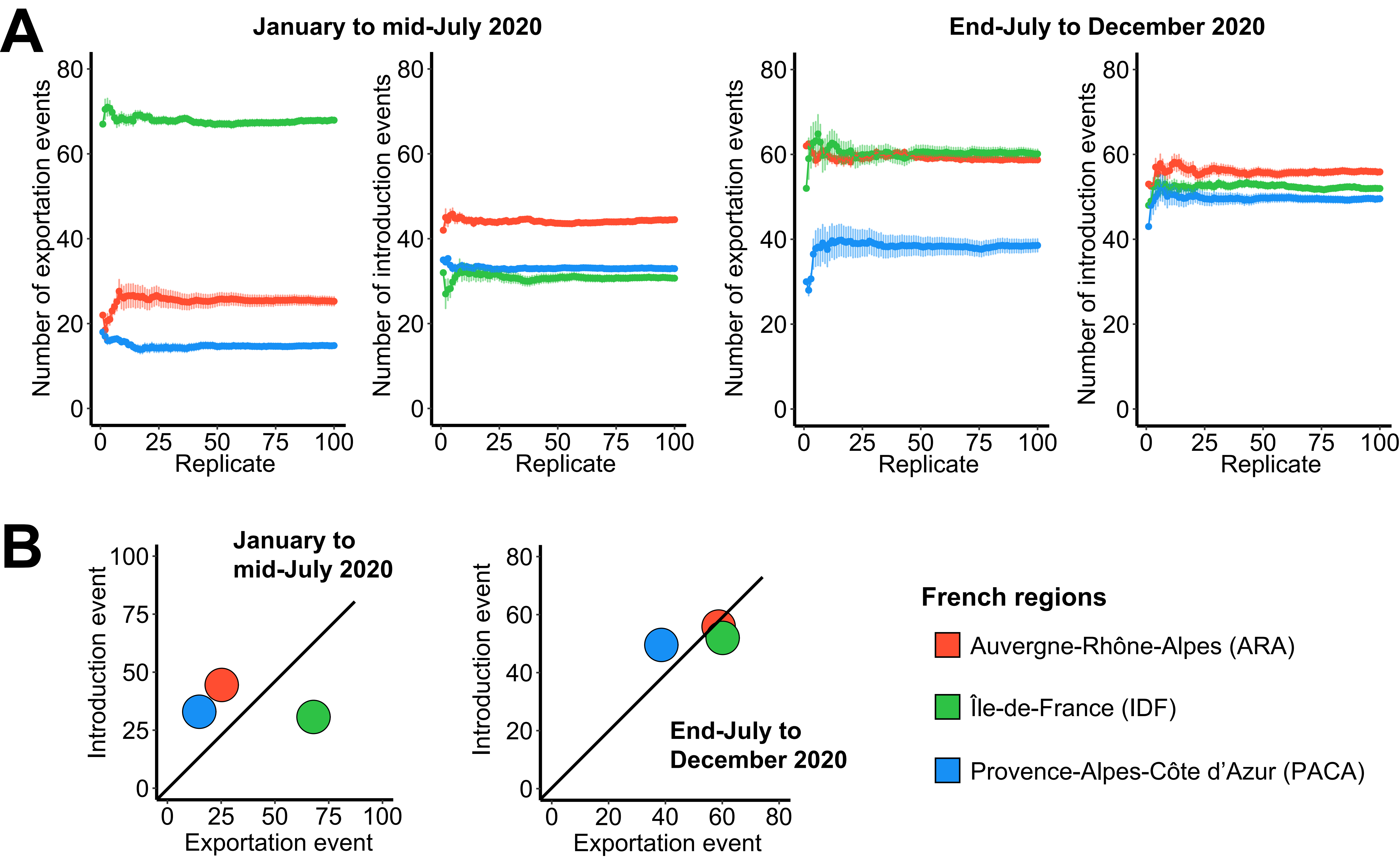

### figure_6-supplement_fig_1.pdf

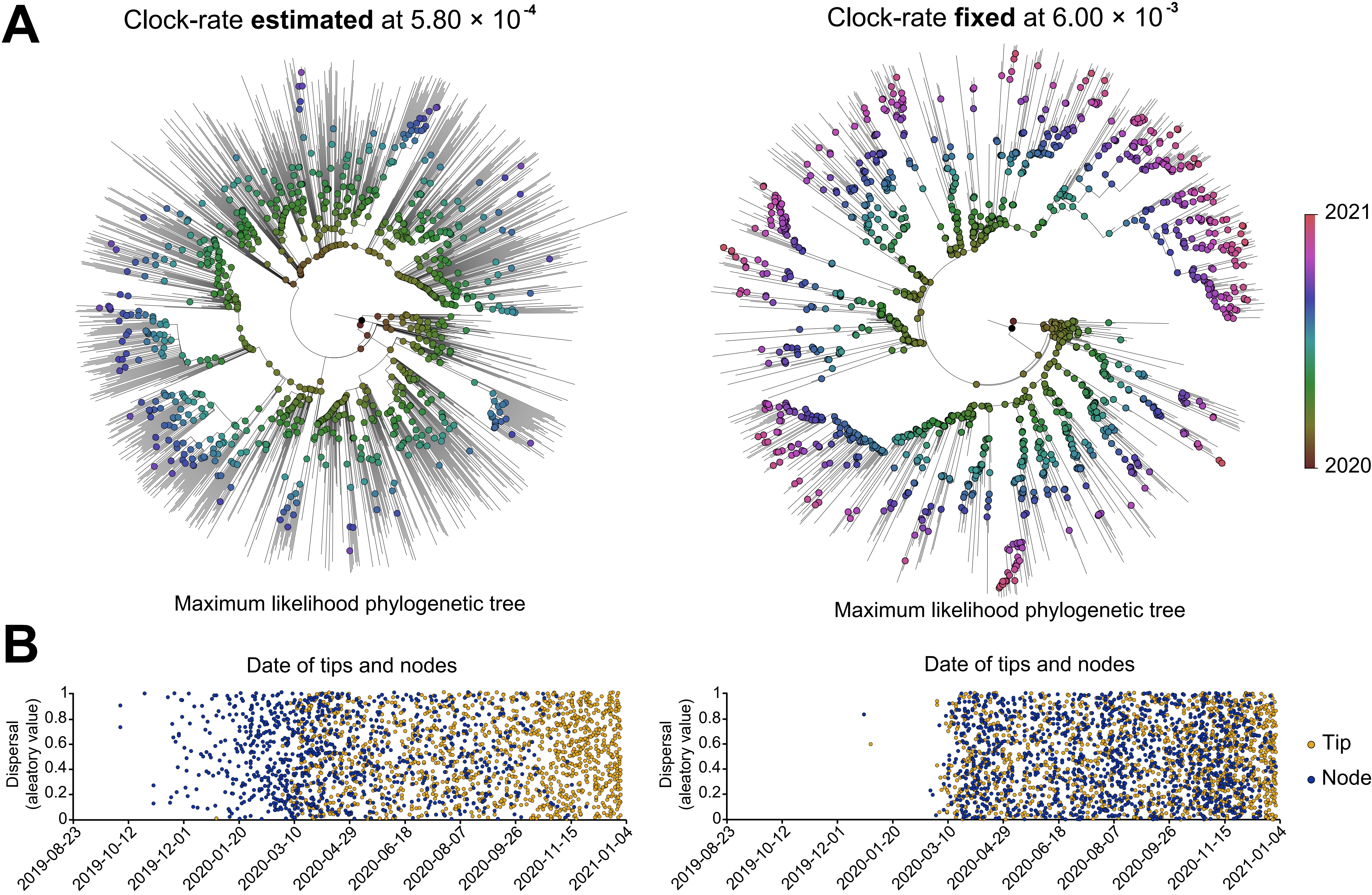

### figure_6-supplement_fig_2.pdf

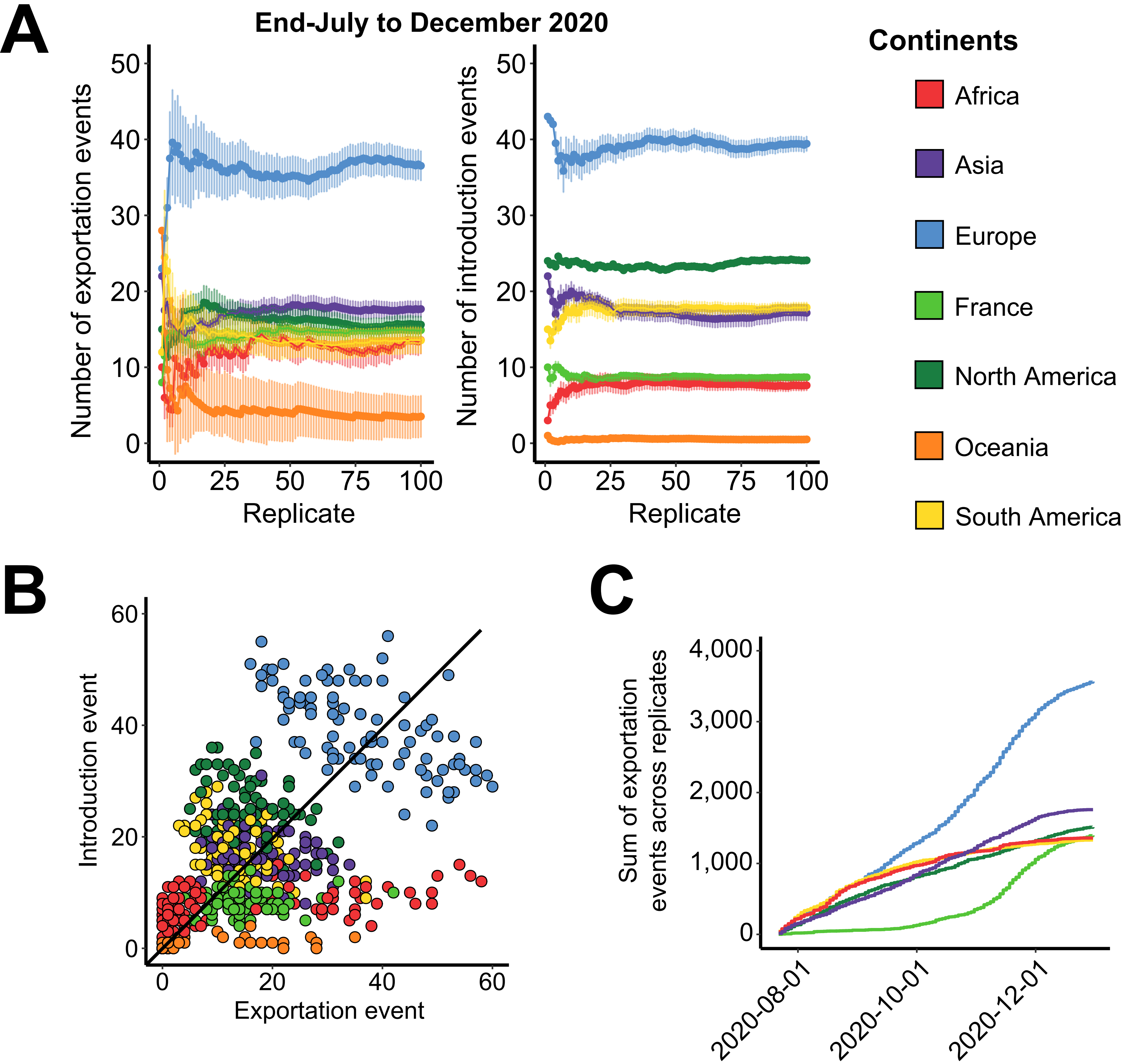
